# MACHINE LEARNING PREDICTION FOR COVID 19 PANDEMIC IN INDIA

**DOI:** 10.1101/2020.05.20.20107847

**Authors:** Roseline Oluwaseun Ogundokun, Joseph Bamidele Awotunde

## Abstract

**Background:** Coronavirus was detected in December 2019 in a bulk seafood shop in Wuhan, China. The original incident of COVID-19 pandemic in India was conveyed on 30th January 2020 instigating from the nation called china. As of 25th April 2020, the Ministry of Health and Family Welfare has established a total of 24, 942 incidents, 5, 210 recuperation including 1 relocation, and 779 demises in the republic.

**Objective:** The objective of the paper is to formulate a simple average aggregated machine learning method to predict the number, size, and length of COVID-19 cases extent and wind-up period crosswise India.

**Method:** This study examined the datasets via the Autoregressive Integrated Moving Average Model (ARIMA). The study also built a simple mean aggregated method established on the performance of 3 regression techniques such as Support Vector Regression (SVR, NN, and LR), Neural Network, and Linear Regression.

**Result:** The results showed that COVID-19 disease can correctly be predicted. The result of the prediction shows that COVID-19 ailment could be conveyed through water and air ecological variables and so preventives measures such as social distancing, wearing of mask and hand gloves, staying at home can help to avert the circulation of the sickness thereby resulting in reduced active cases and even mortality.

**Conclusion:** It was established that the projected method outperformed when likened to previously obtainable practical models on the bases of prediction precision. Hence, putting in place the preventive measures can effectively manage the spread of COVID-19, and also the death rate will be reduced and eventually be over in India and other nations.

## INTRODUCTION

Coronaviruses are a wide intimate of diseases, few of which lead to disease in humans and the remaining which mingle between animals and natures. Animal coronaviruses will occasionally transmit to individuals and only transmit to humans [1]. In recent years, zoonotic coronaviruses have formed triggering humanoid outbursts for instance coronavirus ailment 2019 (COVID-19), severe acute respiratory syndrome (SARS), as well as a respiratory syndrome in the Middle East (MERS). The human disease occurs often as a lung infection, or occasionally as a stomach infection. The clinical range of disease ranges from no signs or moderate breathing problems to extreme, increasingly progressing pneumonia, severe breathing suffering condition, infected tremor, or death-induced multiple-body part catastrophe [2].

As of 25 April 2020, India registered more than 24, 942 established crisis of COVID-19). Around 5, 210 individuals are now in good health from these, whereas 779 crises have led to death. The sum of individuals afflicted with the disease was increasing in the South Asian nation which led to the government swinging into action to further curb the outbreak’s spread. As of mid-April 2020, more than two million cases of coronavirus have been identified worldwide.

A Sikh preacher who, bearing the virus, returned from traveling to Italy and Germany, became a “mega propagator”. He was present at a Sikh commemoration in Anandpur Sahib from 10th to 12th of March 2020 [2, 3]. 27 cases of COVID-19 were backtracked in the direction of him [4]. On 27 March more than 40,000 residents were quarantined in 20 villages in Punjab to control the circulation of the disease [3, 5]. On 31st of March, an occasion of a spiritual assembled group of people in Tablighi Jamaat that happened in Delhi at the beginning of March arose as a novel disease asperity next multiple crisis around the world were backtracked toward the occasion [6]. More than 9 thousand proselytizers might have joined the service, accompanying the bulk arriving from different nations in India [7, 8] including nine hundred and sixty from forty republics abroad [9]. As reported by the Office of Health and Family Wellbeing, this incident was related to 4,291 out of 14,378 confirmed cases in twenty-three Indian cities including unified terrains before the 18th of April 2020 [10]. On the 6th of April 2020, 26 nurses and 3 physicians were found to have been diagnosed with the virus in Mumbai’s Wockhardt Hospital. Hospitals were locked down immediately, and a safety region was professed. Health treatment incompetence was responsible for the infections [11]. In India, COVID-19 infection rates are estimated towards 1.7, slightly lesser than in the worst-pretentious nations [12]. The outburst had proclaimed a widespread in over 12 nations as well as unification terrains, where requirements of the 1897 Infectious Ailments Act were applied, and public institutions and other business enterprises were on standstill. Indian had revoked entirely every traveler entry permit since most reported cases have been related to other countries [13]. On 22 March 2020, at Prime Minister Narendra Modi’s say, India implemented a 14-aera community restriction. The government responded with movement restriction in 75 areas wherever COVID-19 events and all major cities had taken place [14, 15]. Also, the Prime Minister ordered a 21-day national shutdown on 24 March, affecting India’s entire 1.3 billion population [16, 17]. The Prime Minister extended the current national lockout until 3 May on 14 April [18]. Michael Ryan, Chief Executive Officer of the Healthiness Emergencies Program of the World Health Organization, said that India has “great potential” to cope with COVID-19 epidemic as per the next most populated nation, would possess a significant effect proceeding the ability of the globe to cope with the disease [19]. Many analysts were apprehensive concerning the financial destruction instigated via the lockout, having immense consequences on migrant employees, large as well as trivial initiatives, agriculturalists, and entrepreneurial individuals, who were abandoned without an income in the nonappearance of transport and consumer contact [20, 21]. Spectators noted the lockout reduced the pandemic’s progress frequency by 6 April to double every six days [22] and by 18 April to double every eight days [23]. Founded on statistics from seventy-three nations, the Oxford Covid-19 Régime Response Tracker (OxCGRT) states that the Indian Government has reacted to the pandemic more rigorously than other nations. This acknowledged the fast intervention of the government, emergency policymaking emergency healthcare spending, budgetary initiatives, expenditure in vaccine development and successful reaction towards the crisis, and rated India with a “100” aimed at her firmness [24, 25]. India registered the first COVID-19 case in Kerala on 30 January, which grew to 3 incidences by 3rd February 2020; altogether were undergraduates who came back from Wuhan, China [26, 27]. The remainder of February showed no noticeable increase in events. On the 4th of March 2020, twenty-two fresh incidence was discovered together with those groups of visitors from Italy involving 14 of the participants contaminated [28]. The spread intensified in mid-March, amid news of multiple cases around the world, several of which were related to individuals with itinerant antiquity to pretentious nations. On the 12th of March, 2020 an individual of age 76 who had come back from Saudi Arabia turned out to be the country’s primary survivor of the COVID-19 disease [29]. Confirmed deaths reached a hundred on 15th March 2020, [30] 1thousand by 28th March 2020, [31] 5 thousand by 7th of April 2020, [32] 10 thousand by 14th April 2020 [33]. Demise toll passed 50 by 1st April [34] then 100 by 5th April [35].

With COVID-19 no additional antiviral therapy is recommended. Infected patients should provide medical medication to help in pain relief. In extreme cases, the function of vital organs should be protected. [24] SARS-CoV-2 is currently not available as a vaccine. Evitement is the primary form of deterrence. Numerous international projects have quickly arisen to identify and evaluate the efficacy of antivirals, immunotherapies, monoclonal antibodies, and vaccines. Pharmacotherapy Protocols and Reviews for COVID-19 have been written. [25, 36-38].

Transmission of infectious disease is a dynamic mechanism of transmission that happens within the crowd. Frameworks can be developed for this method to potentially examine and test the propagation mechanism of infectious diseases [39] so that we can forecast correctly the future pattern of infectious diseases [40]. Therefore, to monitor or reduce the damage of infectious diseases, the study and review of predictive models for infectious diseases have been a hot topic of science [41].

Therefore, the study projected a simple average aggregated scheme, and the aggregated system has been established by aggregating three regression methods which include Support Vector Regression, Linear Regression as well as Artificial Neural Network. The variables utilized for the formulation of an aggregated method are the figures of COVID-19 cases. The dataset was gathered and collated from Statistica.com from January to April 2020 gathered monthly. The study predicted the values from the previous COVID-19 incidents and values of environment variables such as Water and Air.

## MATERIAL AND METHODOLOGY

### Data Gathering

The authentic datasets of COVID-19 have been gathered from https://www.mygov.in/ and https://www.pharmaceutical-technology.com/ the dataset is publicly available on cases from India from the first case index on January 30 2020. The datasets gathered were in a monthly form that is January 2020 to April 2020. Table 1 displays the scenario of COVID-19 incidents in India from January 2020 to April 2020. As at 25th April 2020 COVID-19 dataset includes accumulated 408, 658 total samples, confirmed cases of 24, 942, recovered cases of 5, 209; 779 death cases and 1 migration.

**Table 1:** Datasets for COVID-19 Cases in India

| Total Samples | Confirmed | Active Cases | Recovered/Discharged Cases | Deceased Cases | Migrated |
| --- | --- | --- | --- | --- | --- |
| 408, 658 | 24, 942 | 18, 953 | 5, 209 | 779 | 1 |

**Figure 1.**
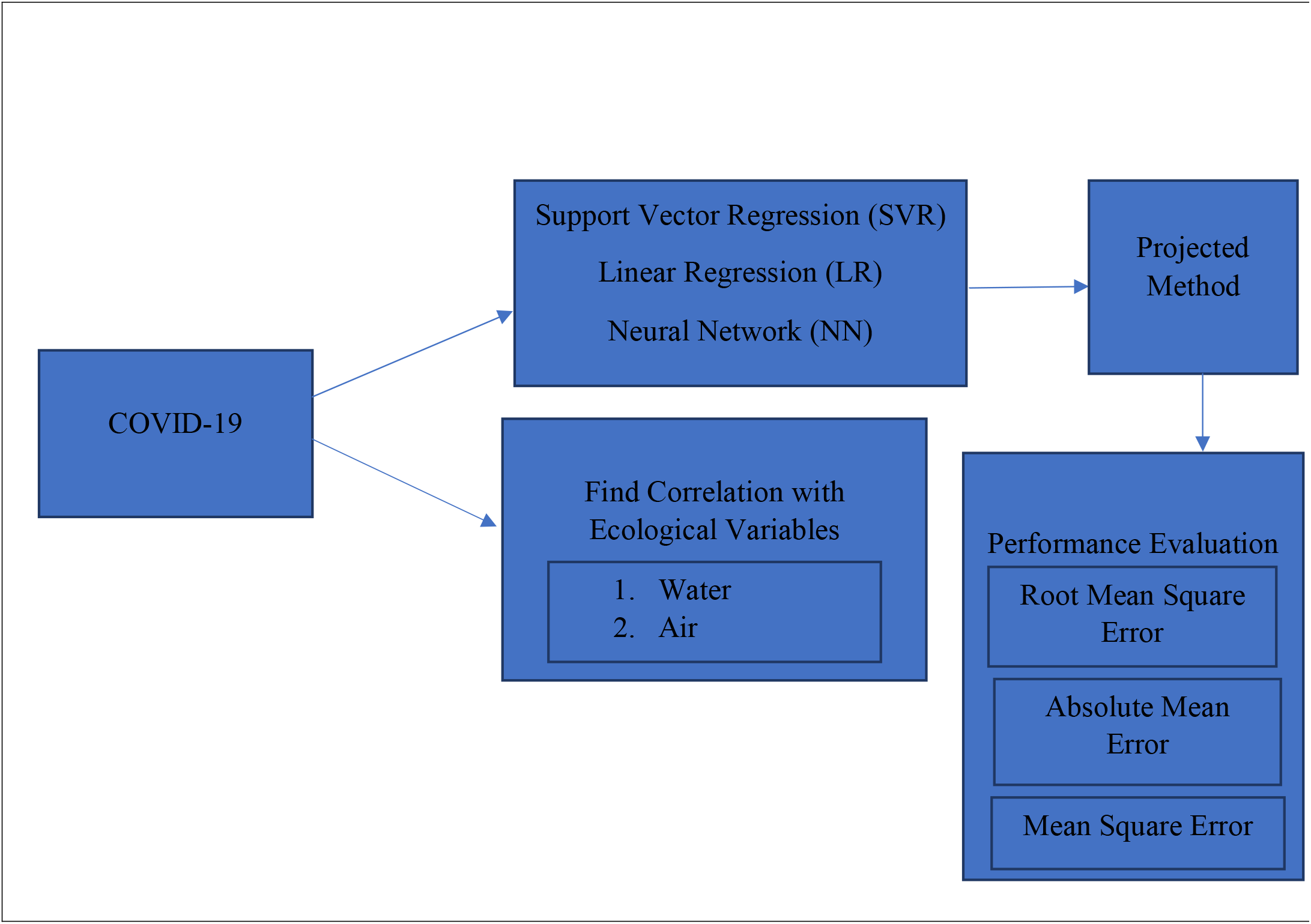
Flow Diagram of Projected Method

### Method Applied

#### Machine Learning Methods

There are numerous diverse methods utilized to perform machine learning tasks. Machine learning approaches require certain types of algorithmic approaches. According to Dataquest, 2020, there are three types of machine learning algorithms and they are:

i. Supervised learning algorithms such as Classification, Regression, and Ensemble
ii. Unsupervised learning algorithm such as Association, Clustering, Dimensionality reduction
iii. Reinforcement learning

##### a. Artificial Neural Network (ANN)

The utmost extensively ANNs utilized in the estimation problem is multi-layer perceptions (MLPs), which employs the solitary tiers feed-forward network [43]. This method is categorized by a system of 3 layers. The nodes in several tiers are as well identified as altering fundamentals.

The outcome of the method is calculated employing the subsequent mathematical expression

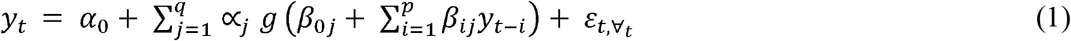

At this point *y_t_*_−_*_i_* (*i* = 1, 2, …… p) are the p entered and *y_t_* is the result. The digits p, q are the figures entered and concealed nodes. ∝*_j_*(*j* = 0, 1, 2, …… q) and *β_ij_*(*i* = 0, 1, 2, …… p); j = 0, 1, 2, …… q) are the formulation weightiness and *ε_t_* is arbitrary tremor; ∝_0_ and *β*_0_*_j_* are the prejudice terms. Generally, the logistics sigmoid function 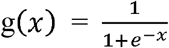 is employed as the nonlinear stimulation function.

##### b. Linear Regression

In statistics, linear regression is the scheme for demonstrating the association amid the scalar or reliant variables and solitary or additional self-determining variables [44, 45, 46, 47, 48]. The circumstance of the descriptive variables is termed a simple linear regression while in the circumstance of greater than single descriptive variables the procedure is referred to as multiple linear regressions. The mathematical formulation of the linear regression is as specified beneath.

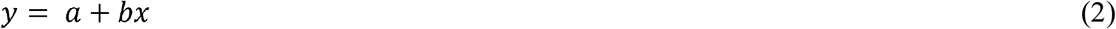

Where *x* and *y* are 2 variables of the regression line, *b* = Grade of the contour and *a* = *Y* interrupt of the contour.

##### c. Support Vector Machines

Support vector machines (SVM) are a supervised learning scheme which could be employed for the classification as well as regression glitches (Adikari et al., 2012; Shanthi et al., 2012). The core impression of SVM once employed to dual categorization difficulties is to discover an established manic level that excellently splits the 2 assumed clusters of tutoring models. In circumstances where data points are not linearly divisible, a lenient border manic level classifier is created as below

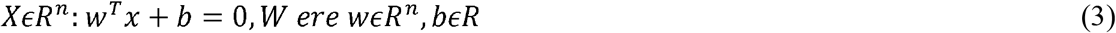

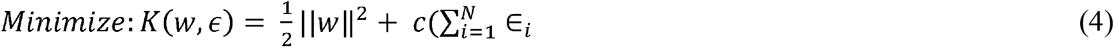

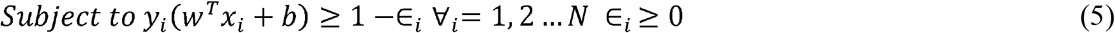

### Support Vector Machine Regression (SVMR)

To employ SVMR, as in the categorization difficulty this belongs to a few clusters, for instance, A1 and A2 in the circumstance of regression and support vector machine at this point is the actual figure and additional variables are equivalent as for the categorization glitches [45]. Regression techniques are one of the prevailing methods for the prediction of specific datasets.

In this study, the authors formulated a simple mean aggregated method by combining 3 popular regression models and predicted the sum of COVID-19 in India. Authors such as [49] and [50] have employed the regression method predominantly to communicable datasets and as well hypotheses an ensemble model typically with 3 estimation approaches.

### Description of Postulated Aggregated Method

There is a key challenge in the study of prediction predominantly in the relation of predicting an occurrence of a specific illness as it had been stated earlier. There are several techniques obtainable in previous research work for the study of forecasting and yet there exists one problem or the other with the performance of the separate technique. To conquer this restraint, the authors postulated an aggregated method. In the study, we designated the method as let 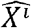 be the group of self-determining variables where 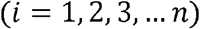. The actual data group of a sequence is demarcated as *X* = [*x*_1_, *x*_2_, − *x_n_* raise to power *T* being its prediction attained from the *i^t^* scheme.

#### Algorithm

**Enter: Group of Independent Variables** 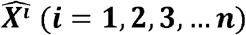.

**Result: Prediction for the Months** *ý*.

1. Employing the independent variables, this revert them to discover the estimation by employing SVR, LR, and ANN
2. Revert figures of entirely the 3 methods and derive a mean of them in a way that *ý* = (*y_l_* + *y*_∝_ + *y_n_*)/3
3. Calculate the predicted figures by the weighted aggregated method
4. Equate the distinct prediction error with a simple mean aggregated method

### Performance Evaluation Metric

The comprehensive examination in this study was achieved in R studio version 3.5. Prediction presentation of entirely the methods was thereafter assessed in the relations of the 3 very recognized errors: Root Mean Square (RMSE), Mean Scaled Error (MSE) and Mean Absolute Error (MAE).

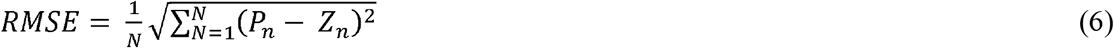

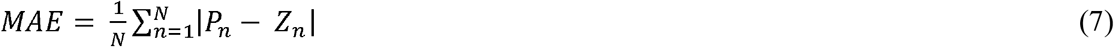

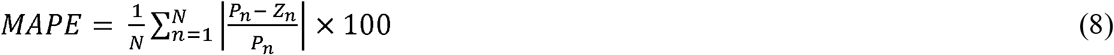

At this point *P_n_* are *Z_n_* real prediction explanations correspondingly; *N* is the overall scope of crises.

## RESULT AND DISCUSSION

### Result Equivalence

This section describes the objective of finding the equivalence between the figures of COVID-19 cases with environmental variables such as water (sewage overflow) and air (air streams or wind). In this study, ecological variables play an important role in the spread of COVID-19 diseases. To accomplish this objective the authors have applied a statistical P-value test to determine the equivalence between the figure of COVID-19 cases. After examination by the P-value test, it was concluded that water is a protuberant ecological variable for the incidence of COVID-19 incident in India. Many authors have found the equivalence between infectious diseases and ecological variables by P-value test [51, 52]. The application of the P-value test accomplishes that ecological variables like water and air had a positive equivalence with the occurrence of several cases for the period since this disease started. The statistical implication was measured p<0.05. The study discards the null hypothesis. The equivalence of COVID-19 cases with water and air is substantial table 2.

**Table 2.**
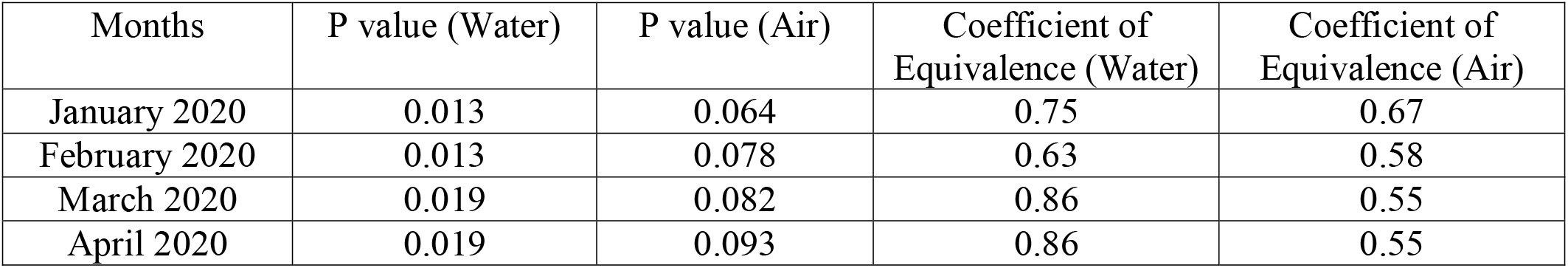
Equivalent Values of COVID-19 with Ecological Variables

This study likewise examined the datasets via the Autoregressive Integrated Moving Average Model (ARIMA). ARIMA is similarly extensively utilized for estimation methodology for the COVID-19 dataset. The study built a simple mean aggregated method established on the performance of 3 regression techniques. It was deduced that the performance of the ARIMA approach wasn’t improved in terms of the error metrics and the best 3 regression scheme for the aggregate as the complication subject is there. This is shown in table 3.

**Table 3.** Performance by ARIMA Technique

| Dataset | RMSE | MSE | MAE |
| --- | --- | --- | --- |
| COVID-19 | 286.879 | 78604.436 | 175.672 |

### Generation of Revert Figures

Table 4 to 6 illustrates the regression figures for all the methods. After employing entirely, the 3 regression methods on the datasets and attained the predicted figure of the pandemic, the mean of the figures are attained as shown in table 7.

**Table 4.** Revert Figures (Support Vector Regression)

| Months | COVID-19 |
| --- | --- |
| 1 | 25.483679 |
| 2 | 89.637427 |
| 3 | 48.246173 |
| 4 | 24.520456 |

**Table 5.** Revert Figures (Linear Regression)

| Months | COVID-19 |
| --- | --- |
| 1 | 185.57382 |
| 2 | 57.69134 |
| 3 | 147.32278 |
| 4 | 186.34657 |

**Table 6.**
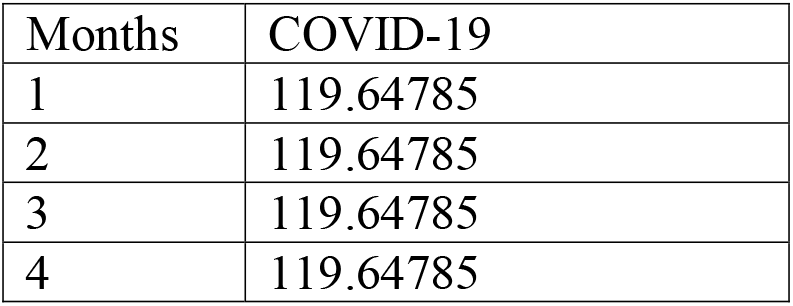
Revert Figures (Neural Network)

| Months | COVID-19 |
| --- | --- |
| 1 | 119.64785 |
| 2 | 119.64785 |
| 3 | 119.64785 |
| 4 | 119.64785 |

**Table 7.** Mean Figures for the entirely COVID-19 Crises

| Months | COVID-19 |
| --- | --- |
| 1 | 109.4234 |
| 2 | 88.6456 |
| 3 | 118.6324 |
| 4 | 109.7459 |

### Evaluation of Prediction Precision

Tables 8-11 displays the evaluation of the prediction precision in terms of root mean square error, mean scaled error and mean absolute error.

**Table 8.**
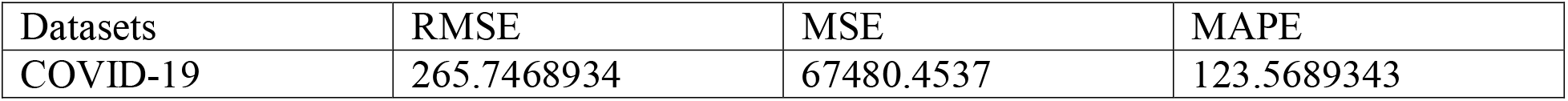
Support Vector Regression

| Datasets | RMSE | MSE | MAPE |
| --- | --- | --- | --- |
| COVID-19 | 265.7468934 | 67480.4537 | 123.5689343 |

**Table 9.** Linear Regression

| Datasets | RMSE | MSE | MAPE |
| --- | --- | --- | --- |
| COVID-19 | 259.46783 | 56777.2316 | 164.674563 |

**Table 10.** Neural Network

| Datasets | RMSE | MSE | MAPE |
| --- | --- | --- | --- |
| COVID-19 | 246.7356982 | 64321.37210 | 161.467839 |

**Table 11.** Postulated Aggregated Method

| Datasets | RMSE | MSE | MAPE |
| --- | --- | --- | --- |
| COVID-19 | 12.245912 | 167.4568321 | 7.678 |

### Outcomes

For the confirmation of our postulated aggregated system, one instantaneous series dataset was utilized in this research. These are the numeral of COVID-19 crises in India and these datasets were gathered on Statistica.com. The explanation of period series datasets is obtainable in Table 12.

**Table 12.** Explanation of Datasets

| Time Series | Description |
| --- | --- |
| COVID-19 | Figure of recorded COVID-19 crises as of January 2020 to April 2020 |

The figures of all these errors are anticipated is to be as low as probable for improved prediction accuracy.

## DISCUSSION

As a substitute for epidemiologic spread procedure, the study employed 3 aggregated methods SVR, NN, and LR to predict the instantaneous movement of the conveyance dynamics and generate the real-time predictions of COVID-19 disease transversely the metropolises of India. The finding presented that the precision of estimation and succeeding multiple-process prediction was extreme. It was revealed that predicting enhanced when aggregating 3 regression models together compared to when used individually. The postulated system appraised the possible period when there will be decreasing evolution of fresh established cases crosswise the metropolises and the extent of COVID-19 disease across India and the minute the figure of the accrued established incident of COVID-19 would spread to the upland of their accrued crises.

The core intention of this investigation is to formulate a simple mean aggregated method for the estimation of COVID-19 disease in India. To formulate this, the study first computed the revert figures of the COVID-19 disease from the whole separate regression procedure followed by their amalgamation in an aggregated method. In the second stage of formulating the method, the study intention is to decrease the prediction errors of COVID-19 in India and these errors include RMSE, MAE, and MAPE. Each time a certain method gives a greater prediction error compared to alternative methods its weightiness in the aggregate is diminished and vice versa. This study has joined the 3 employed methods and presented the data for COVID-19 disease in India as stated below. In this study, the formulation of aggregated methods illustrates a substantial enhancement in the prediction of the COVID-19 disease in India. The postulated aggregated method is likened with the separate prediction of these methods. The attained COVID-19 prediction precision attained for the entire approach is represented in the table 8-10. The obtainable outcome of COVID-19 disease is shown in Table 13 which demonstrates that our postulated aggregated system delivered the least prediction error amid the entire separate fitted methods. The study delivered a substantial enhancement in prediction precisions for COVID-19 disease in India when the postulated aggregated system was employed.

**Table 13.**
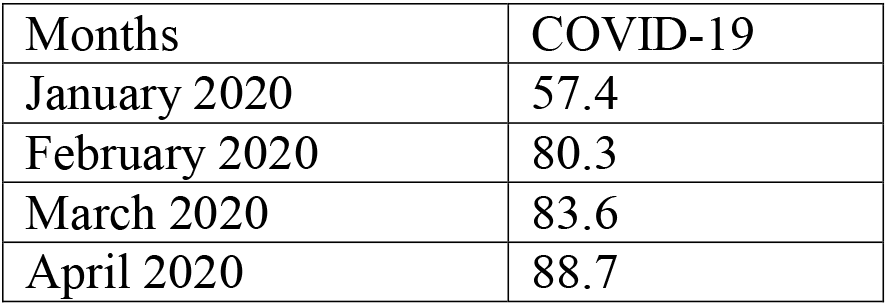
Prediction Table from Postulated Method

If the datasets are dependable and there exist no subsequent outbreak, the aggregated methods predicted that COVID-19 outburst in India could end by May ending. The aggregated methods permit entering the interferences information as well as examining the influence of interferences on the extent of the disease outburst and the ending period if the COVID-19 epidemic. Employing the assumed original extents of the COVID-19 outburst, the study utilized the aggregated methods (SVR, NN, and LR) with identified framework and model to evaluate the extent of the outburst in time to come and excite the influence of the interferences on the magnitude and asperity of the pandemic. Approbatory to the individually used methods for the conveyance of the COVID-19 diseases, the aggregated methods (SVR, NN, and LR) provide real-time predicting instruments used for shaping and tracking COVID-19 disease in India, reckoning the COVID-19 disease, obtaining COVID-19 disease asperity, predicting the extent of the pandemic together with supporting government and health staffs to constitute strategy and competent verdicts towards the eradication of the COVID-19 diseases in India.

## CONCLUSION

Approbatory to the individually used methods for the conveyance of the COVID-19 diseases, the aggregated methods (SVR, NN, and LR) provide real-time predicting instruments used for shaping and tracking COVID-19 disease in India, reckoning the COVID-19 disease, obtaining COVID-19 disease asperity, predicting the extent of the pandemic together with supporting government and health staffs to constitute strategy and competent verdicts towards the eradication of the COVID-19 diseases in India.

The integration of the prediction from diverse methods substantially decreases prediction errors and consequently makes available advanced precision. A few decades back, several researchers’ studies have suggested several statistical methods. The study postulated a simple-mean aggregated method for the prediction of COVID-19 disease in India. The projected aggregated approach investigated separate predictions and therefore the prediction of COVID-19 disease in India is formulated with 3 very recognized methods: Neural Network, Support Vector Regression, and Linear Regression. The result indicates that the postulated method outperformed based on prediction precision of the COVID-19 disease in India when likened to currently existing applicable methods.

## FUTURE WORK

In the future the competence of the postulated method could be as well as be investigated and some other regression models or algorithms can be used and evaluated.

## Data Availability

The authentic datasets of COVID-19 have been gathered from https://www.mygov.in/ and https://www.pharmaceutical-technology.com/ the dataset is publicly available on cases from India from the first case index on January 30, 2020

https://www.pharmaceutical-technology.com/

## REFERENCES

1. BMJBest Practice, 2020. Overview of Coronaviruses. https://bestpractice.bmi.com/topics/en-us/3000165

2. Smith JD, MacDougall CC, Johnstone J, Copes RA, Schwartz B, Garber GE. Effectiveness of N95 respirators versus surgical masks in protecting health care workers from acute respiratory infection: a systematic review and meta-analysis. CMAJ. 2016 May 17. 188 (8):567–574. [Medline].

3. Bae S, Kim MC, Kim JY, Cha HH, Lim JS, Jung J, et al. Effectiveness of Surgical and Cotton Masks in Blocking SARS-CoV-2: A Controlled Comparison in 4 Patients. Ann Intern Med. 2020 Apr 6. [Medline].

4. World Health Organization. Coronavirus disease 2019 (COVID-19) Situation Report—48. World Health Organization. Available at https://www.who.int/docs/default-source/coronaviruse/situation-reports/20200308-sitrep-48-covid-19.pdf?sfvrsn=16f7ccef_4. March 8, 2020; Accessed: March 9, 2020.

5. CDC. 2019 Novel Coronavirus, Wuhan, China: 2019 Novel Coronavirus (2019-nCoV) in the U.S. Centers for Disease Control and Prevention (CDC). Available at https://www.cdc.gov/coronavirus/2019-ncov/cases-in-us.html. March 18, 2020; Accessed: March 19, 2020.

6. Otto MA. Wuhan Virus: What Clinicians Need to Know. Medscape Medical News. Available at https://www.medscape.com/viewarticle/924268. January 27, 2020; Accessed: January 27, 2020.

7. Feuer W, Higgins-Dunn N, Lovelace B. US now has more coronavirus cases than either China or Italy. CNBC. Available at https://www.cnbc.com/2020/03/26/usa-now-has-more-coronavirus-cases-than-either-china-or-italy.html. March 26, 2020; Accessed: March 27, 2020.

8. Severe Outcomes Among Patients with Coronavirus Disease 2019 (COVID-19) — United States, February 12–March 16, 2020. MMWR Morb Mortal Wkly Rep. 2020 Mar 18. 69:[Full Text].

9. Bernstein L, McGinley L, Sun LH. Northern California coronavirus patient wasn’t tested for days. The Washington Post. Available at https://www.washingtonpost.com/health/northern-californian-tests-positive-for-coronavirus-in-first-us-case-with-no-link-to-foreign-travel/2020/02/26/b2088840-58fb-11ea-9000-f3cffee23036_story.html. February 27, 2020; Accessed: February 27, 2020.

10. Li Q, Guan X, Wu P, Wang X, et al. Early Transmission Dynamics in Wuhan, China, of Novel Coronavirus-Infected Pneumonia. N Engl J Med. 2020 Jan 29. [Medline].

11. Chan JF, Yuan S, Kok KH, To KK, Chu H, Yang J, et al. A familial cluster of pneumonia associated with the 2019 novel coronavirus indicating person-to-person transmission: a study of a family cluster. Lancet. 2020 Jan 24. [Medline].

12. CDC. Coronavirus Disease 2019 (COVID-19): COVID-19 Situation Summary. CDC. Available at https://www.cdc.gov/coronavirus/2019-ncov/summary.html. February 29, 2020; Accessed: March 2, 2020.

13. Ferguson NM, Laydon D, Nedjati-Gilani G, Imai N, Ainslie K, Baguelin M, et al. Impact of non-pharmaceutical interventions (NPIs) to reduce COVID-19 mortality and healthcare demand. Imperial College COVID-19 Response Team. Available at https://www.imperial.ac.uk/media/imperial-college/medicine/sph/ide/gida-fellowships/Imperial-College-COVID19-NPI-modelling-16-03-2020.pdf. 2020 Mar 16; Accessed: March 18, 2020.

14. Brunk D. CDC: First Person-to-Person Spread of Novel Coronavirus in US. Medscape Medical News. Available at https://www.medscape.com/viewarticle/924571. January 30, 2020; Accessed: January 31, 2020.

15. U.S. reports its first case of person-to-person transmission. The New York Times. Available at https://www.nytimes.com/2020/01/30/world/asia/coronavirus-china.html#link-69c13d84. January 30, 2020; Accessed: January 30, 2020.

16. Centers for Disease Control and Prevention. Coronavirus Disease 2019 (COVID-19): People at Higher Risk. Centers for Disease Control and Prevention. Available at https://www.cdc.gov/coronavirus/2019-ncov/specific-groups/high-risk-complications.html. March 8, 2020; Accessed: March 9, 2020.

17. CDC. 2019 Novel Coronavirus, Wuhan, China: Symptoms. CDC. Available at https://www.cdc.gov/coronavirus/2019-ncov/about/symptoms.html. January 26, 2020; Accessed: January 27, 2020.

18. Lauer SA, Grantz KH, Bi Q, Jones FK, Zheng Q, Meredith HR, et al. The Incubation Period of Coronavirus Disease 2019 (COVID-19) From Publicly Reported Confirmed Cases: Estimation and Application. Ann Intern Med. 2020 Mar 10. [Medline].

19. Wu Z, McGoogan JM. Characteristics of and Important Lessons From the Coronavirus Disease 2019 (COVID-19) Outbreak in China: Summary of a Report of 72 - 314 Cases From the Chinese Center for Disease Control and Prevention. JAMA. 2020 Feb 24. [Medline].

20. Rabin RC. Lost Sense of Smell May Be Peculiar Clue to Coronavirus Infection. The New York Times. Available at https://www.nytimes.com/2020/03/22/health/coronavirus-symptoms-smell-taste.html. March 22, 2020; Accessed: March 24, 2020.

21. Dong Y, Mo X, Hu Y, Qi X, Jiang F, Jiang Z, et al. Epidemiological Characteristics of 2143 Pediatric Patients With 2019 Coronavirus Disease in China. Pediatrics. 2020 Mar 16. [Medline].

22. Qiu H, Wu J, Hong L, Luo Y, Song Q, Chen D. Clinical and epidemiological features of 36 children with coronavirus disease 2019 (COVID-19) in Zhejiang, China: an observational cohort study. Lancet Infect Dis. 2020 Mar 25. [Medline].

23. CDC. 2019 Novel Coronavirus, Wuhan, China: Interim Healthcare Infection Prevention and Control Recommendations for Patients Under Investigation for 2019 Novel Coronavirus. CDC. Available at https://www.cdc.gov/coronavirus/2019-ncov/infection-control.html. January 18, 2020; Accessed: January 27, 2020.

24. CDC. 2019 Novel Coronavirus, Wuhan, China: Prevention & Treatment. CDC. Available at https://www.cdc.gov/coronavirus/2019-ncov/about/prevention-treatment.html. January 26, 2020; Accessed: January 27, 2020.

25. McCreary EK, Pogue JM. COVID-19 Treatment: A Review of Early and Emerging Options. Open Forum Infectious Diseases (OFID). 2020 Mar 23. [Full Text].

26. CDC. 2019 Novel Coronavirus, Wuhan, China: Frequently Asked Questions and Answers. CDC. Available at https://www.cdc.gov/coronavirus/2019-ncov/faq.html. January 27, 2020; Accessed: January 27, 2020.

27. van Doremalen N, Bushmaker T, Morris DH, Holbrook MG, Gamble A, Williamson BN, et al. Aerosol and Surface Stability of SARS-CoV-2 as Compared with SARS-CoV-1. N Engl J Med. 2020 Mar 17. [Medline].

28. Chin AWH, Chu JTS, Perera MRA, et al. Stability of SARS-CoV-2 in different environmental conditions. The Lancet Microbe. April 2, 2020. [Full Text].

29. Wölfel R, Corman VM, Guggemos W, Seilmaier M, Zange S, Müller MA, et al. Virological assessment of hospitalized patients with COVID-2019. Nature. 2020 Apr 1. [Medline].

30. Zhou F, Yu T, Du R, Fan G, Liu Y, Liu Z, et al. Clinical course and risk factors for mortality of adult inpatients with COVID-19 in Wuhan, China: a retrospective cohort study. Lancet. 2020 Mar 11. [Medline].

31. Liu Y, Yan LM, Wan L, Xiang TX, Le A, Liu JM, et al. Viral dynamics in mild and severe cases of COVID-19. Lancet Infect Dis. 2020 Mar 19. [Medline].

32. Bai Y, Yao L, Wei T, Tian F, Jin DY, Chen L, et al. Presumed Asymptomatic Carrier Transmission of COVID-19. JAMA. 2020 Feb 21. [Medline].

33. Yu P, Zhu J, Zhang Z, Han Y, Huang L. A familial cluster of infection associated with the 2019 novel coronavirus indicating potential person-to-person transmission during the incubation period. J Infect Dis. 2020 Feb 18. [Medline].

34. Zou L, Ruan F, Huang M, Liang L, Huang H, Hong Z, et al. SARS-CoV-2 Viral Load in Upper Respiratory Specimens of Infected Patients. N Engl J Med. 2020 Feb 19. [Medline].

35. Leung NHL, Chu DKW, Shiu EYC, et al. Respiratory virus shedding in exhaled breath and efficacy of face masks. Nat Med. 2020. [Full Text].

36. [Guideline] Bhimraj A, Morgan RL, Shumaker AH, et al. Infectious Diseases Society of America Guidelines on the Treatment and Management of Patients with COVID-19. IDSA. Available at https://www.idsociety.org/practice-guideline/covid-19-guideline-treatment-and-management/. April 13, 2020; Accessed: April 13, 2020.

37. Sanders JM, Monogue ML, Jodlowski TZ, Cutrell JB. Pharmacologic Treatments for Coronavirus Disease 2019 (COVID-19): A Review. JAMA. 2020 Apr 13. [Medline]. [Full Text].

38. Barlow A, Landolf KM, Barlow B, Yeung SYA, Heavner JJ, Claassen CW, et al. Review of Emerging Pharmacotherapy for the Treatment of Coronavirus Disease 2019. Pharmacotherapy. 2020 Apr 7. [Medline]. [Full Text].

39. Kucharski A, Russell T, Diamond C, Liu Y, CMMID nCoV working group, Edmunds J, Funk S, Eggo R. Analysis and projections of transmission dynamics of nCoV in Wuhan. 2020. https://cmmid.github.io/ncov/wuhan_early_dynamics/index.html.

40. Tuite AR, Fisman DN. Reporting, epidemic growth, and reproduction numbers for the 2019 novel coronavirus (2019-nCoV) epidemic. Ann Intern Med. 2020 Feb 5. doi: 10.7326/M20-0358. [Epub ahead of print].

41. Funk S, Camacho A, Kucharski AJ, Eggo RM, Edmunds WJ. Real-time forecasting of infectious disease dynamics with a stochastic semi-mechanistic model. Epidemics 2018; 22: 56–61.

42. Dataquest, 2020. Retrieved on 24^th^ April 2020 from https://community.dataquest.io/c/social/covid19

43. Adhikari, R. & Agrawal, R.K. (2012). Forecasting strong seasonal time series with artificial neural networks. Journal of Scientific and Industrial Research 10: 657–666.

44. Science, C. (2003). Dimensionality Reduction for Indexing Time Series Based on the Minimum Distance Work 711: 697–711.

45. Yang, S.H., Liang, C.K. & Hsieh, C.Y. (2005). Watermarking MPEG-4 2D mesh animation with time-series analysis. Journal of Information Science and Engineering 21: 341–359.

46. Barrow, D.K., Crone, S.F. & Kourentzes, N. (2010). An evaluation of neural network ensembles and model selection for time series prediction. The 2010 International Joint Conference on Neural Networks (IJCNN), 1–8.

47. Shanthi, S. & Kumar, D. (2012). Prediction of blood glucose concentration ahead of time with feature based neural network. Malaysian Journal of Computer Science 25: 136–148.

48. Shaikh, M.S. & Dote, Y. (1999). An Empirical Application of Linear Regression Method and Fir Network for Fault 12: 57–63.

49. Yamana, T.K., Kandula, S. & Shaman, J. (2017). Individual versus super ensemble forecasts of seasonal influenza outbreaks in the United States. PLoS Comput Biol 13: e1005801.

50. Buczak, A.L., Baugher, B., Moniz, L.J., Bagley, T., Babin, S.M. & Guven, E. (2018). Ensemble method for dengue prediction. PLoS ONE 13: e0189988

51. Potts, J.A., Thomas, S.J. & Srikiatkhachorn. (2010). Classification of dengue illness based on readily available laboratory data. American Journal of Tropical Medicine and Hygiene 83: 781–788.

52. Phakhounthong, K., Chaovalit, P. & Jittamala, P. (2018). Predicting the severity of dengue fever in children on admission based on clinical features and laboratory indicators: Application of classification tree analysis. BMC Pediatrics 18: 1–9.

